# Associations between early-life socioeconomic, family and child factors, and anxiety and depression co-developmental trajectories in Brazil - analysis of the 2004 Pelotas Birth Cohort

**DOI:** 10.64898/2026.09.07.26362409

**Authors:** Lucy Barrass, Bushra Farooq, Lucy Riglin, Bryony Weavers, Ana Goncalves Soares, Hannah Jones, Ian Penton-Voak, Luciana Tovo-Rodrigues, Iná S. Santos, Nanette R Lee, Frances Rice, Nicky Lidbetter, Jon Heron, Duleeka Knipe, Laura Howe, Alicia Matijasevich, Amy Shakeshaft

**Affiliations:** Population Health Sciences, Bristol Medical School, University of Bristol, Bristol, UK; The Wolfson Centre for Young People’s Mental Health, Division of Psychological Medicine and Clinical Neurosciences, Cardiff University, Cardiff, UK; Centre for Neuropsychiatric Genetics and Genomics, Division of Psychological Medicine and Clinical Neurosciences, Cardiff University, Cardiff, UK; School of Psychological Science, University of Bristol, Bristol, UK; NIHR Biomedical Research Centre at the University Hospitals Bristol NHS Foundation Trust, Bristol, UK; Postgraduate Program in Epidemiology, Federal University of Pelotas, Pelotas, Rio Grande do Sul, Brazil; Office of Population Studies Foundation, University of San Carlos, Cebu City, Philippines; Anxiety UK, Manchester, UK; South Asian Clinical Toxicology Research Collaboration, Faculty of Medicine, University of Peradeniya, Peradeniya, Sri Lanka; Departamento de Medicina Preventiva, Faculdade de Medicina FMUSP, Universidade de São Paulo, São Paulo, Brasil

**Author notes:** denotes equal contribution (Alicia Matijasevich, Amy Shakeshaft).

## Abstract

**Background:** Anxiety and depression often co-occur and typically arise early in the life course. Despite the high prevalence of these conditions in low- and middle-income countries, evidence of early-life factors associated with their developmental patterns remains limited in these settings. Understanding these factors could shape prevention efforts.

**Methods:** Using the Brazil-based Pelotas 2004 Birth Cohort (n=3,815), we examined associations between socioeconomic, maternal, and child factors and previously-derived co-developmental trajectories of anxiety and depression across childhood and adolescence. The likelihood of anxiety and depression diagnoses was assessed using the Development and Wellbeing Assessment bands at ages 6, 11 and 15. Early-life factors were measured from birth to age 6.

**Results:** Previous analyses identified four anxiety and depression co-development trajectories across childhood and adolescence, characterised by stable-low (53.9%), increasing (28.1%), decreasing (15.1%) and persistent-high (2.9%) likelihood of diagnoses. Using the stable-low class as the reference, maternal depression (child aged 1) and childhood emotional/behavioural problems (age 4) were associated with all three other classes (odds ratio (OR) range for maternal depression: 2.52-5.49; for emotional/behavioural problems: 1.06-1.10). Childhood trauma was associated with decreasing (OR: 3.62, 95% CI: 2.44, 5.37) and persistent-high (OR: 3.21, 95% CI: 1.51, 6.85) classes. Low maternal education (OR: 2.90, 95% CI: 1.28, 6.54) and low support from the child’s father during pregnancy (OR: 2.77, 95% CI: 1.20, 6.42) were associated with the persistent-high class. Maternal cohabitation with a partner at birth was associated with lower odds of children being in the increasing class (OR: 0.65, 95% CI: 0.44, 0.96).

**Conclusion:** The co-development of anxiety and depression showed associations with a range of early-life factors. Some factors were associated with anxiety and depression regardless of age at onset (e.g., maternal depression), whilst others were more strongly associated with adolescent-onset (e.g., maternal cohabitation) or persistence across both developmental periods (e.g., maternal education).

**What’s known?:**

- Mental disorders have been recognized as a major contributor to global disease burden, exhibiting high prevalence among younger populations.
- Anxiety and depression often co-occur. However, little is known about the early-life factors associated with co-occurrence across childhood and adolescence, particularly in low- and middle-income countries (LMICs).

**What’s new?:**

- Using a Brazilian longitudinal birth cohort, we identified socioeconomic, maternal and child factors associated with anxiety and depression co-developmental trajectories.
- Some factors showed associations across both childhood and adolescence (e.g., maternal depression) whilst others were more specific to childhood-onset (e.g., trauma) or adolescent-onset (e.g., maternal cohabiting).

**What’s relevant?:**

- Our findings identify early-life risk factors associated with co-development of anxiety and depression in an LMIC setting, highlighting potential targets for interventions to prevent adverse trajectories.

## Introduction

Mental health conditions are a major public health concern and a leading cause of disease-related burden worldwide, particularly in childhood and adolescence, where rates of depression and anxiety are rapidly increasing (Dongjun et al., 2025; Kieling et al., 2024). Anxiety and depression are two of the most common mental health conditions and often develop early in life. Anxiety often onsets in childhood, whereas depression typically onsets between adolescence and young adulthood (Rapee et al., 2009; Solmi et al., 2022). Anxiety and depression co-occurrence is associated with worse outcomes, including higher rates of suicidal behaviours, and more severe symptoms and clinical course, compared to experiencing either on its own (Fava et al., 2004; Fava et al., 2008; Kessler et al., 2015; Seo et al., 2011).

Understanding the developmental course of these two conditions, and the heterogeneity of development in the population, can help identify groups at highest risk of mental health difficulties that persist into adult life. Little is known about differential patterns of anxiety and depression co-development, particularly in low- and middle-income countries (LMICs) where the burden of these conditions is greater. Our recent work, using data from the 2004 Pelotas Birth Cohort in Brazil, showed four trajectories of anxiety and depression co-development across childhood and adolescence: *stable-low anxiety and depression (53.9%), increasing anxiety and depression (28.1%), decreasing anxiety and depression* (15.1%) *and persistent-high anxiety and depression (2.9%) (Shakeshaft et al., 2026)*. The increasing and persistent-high classes showed elevated odds of later depression and anxiety at age 18, but no increased risk was observed for the decreasing class, indicating higher probability of diagnoses in adolescence may be most predictive of mental health difficulties in young adulthood.

Nevertheless, whilst these trajectories have been identified in Brazil, little is known about the early-life socioeconomic, maternal and child factors associated with membership of these distinct co-developmental trajectories. Examining factors associated with the development of both conditions together may identify common targets for prevention and inform interventions addressing both conditions simultaneously. This is particularly crucial in LMICs, where resources are more limited. Identifying factors associated with the onset and persistence of anxiety and depression in these settings is an important first step towards understanding potential causal mechanisms.

Where there is evidence on factors associated with anxiety and depression co-developmental trajectories, it is exclusively from high-income countries (HIC). These HIC studies have consistently found parental psychopathology, particularly maternal and paternal depression, to be associated with trajectories characterised by elevated or worsening anxiety and depression co-development regardless of age at onset, compared with stable-low trajectories (de Lijster et al., 2019; Olino et al., 2014). Negative parenting behaviours, including harsh punishment and low warmth, have also been shown to be associated with decreasing, increasing and persistent trajectories of anxiety and depression co-development in adolescence (Olino et al., 2014). Ethnic minority groups have shown differing associations by developmental stage, with higher odds of anxiety and depression trajectories characterised by preschool-onset decreasing symptoms, but lower odds of anxiety and depression onset during early to mid-childhood, when compared to the majority ethnic group (de Lijster et al., 2019). Lower maternal education and overall socioeconomic status have been associated with trajectories characterised by elevated, increasing and/or decreasing anxiety and depression symptoms in child and adolescent populations (de Lijster et al., 2019; Songco et al., 2020).

Despite the importance of early childhood as a sensitive period for health and development, relatively few early-life factors associated with anxiety and depression co-developmental trajectories have been examined (Maggi et al., 2010; Walker et al., 2011). Additionally, existing studies have explored anxiety and depression during either childhood or adolescence, rather than across both developmental periods. Evidence examining factors associated with anxiety and depression co-development across childhood and adolescence in LMICs is particularly limited. These associations may differ from those observed in HICs given higher levels of childhood adversity, greater levels of parental mental health difficulties, housing and food insecurity, and barriers to accessing mental health treatment (Rathod et al., 2017; Soares et al., 2016).

Using the Brazilian 2004 Pelotas Birth Cohort, we aimed to explore early-life exposures, including socioeconomic, maternal and child factors, associated with the co-developmental course of anxiety and depression across childhood and adolescence. We hypothesised that lower socioeconomic position, exposure to non-supportive familial environments, worse maternal mental health, and experiencing emotional/behavioural problems or trauma in childhood would be associated with elevated anxiety and depression across development. Further, we hypothesised that these associations may differ based on age of onset of anxiety and depression and persistence across development.

## Methods

An analysis plan was pre-registered on OSF (Open Science Framework, 2025). Deviations from the analysis plan can be found in the transparent change document (Supporting Information).

### Sample

Pelotas is a city in Southern Brazil with a current population of around 326,000 people (Brazilian Institute of Geography and Statistics). Participants for the 2004 Pelotas Cohort were recruited from all births to mothers living in the urban area of Pelotas between 1^st^ January and 31^st^ December 2004. There were 4,263 eligible births and over 99% of mothers agreed to participate (n=4,231) (Santos et al., 2011; Tovo-Rodrigues et al., 2024). Mothers and children have been followed up into adulthood.

### Anxiety and depression trajectory measures

Anxiety and major depressive disorder were assessed using the Development and Well-Being Assessment (DAWBA), a structured clinical interview designed to generate psychiatric diagnoses (Goodman et al., 2000). The DAWBA has been validated in several countries, including Brazil, and several languages, including Portuguese (Fleitlich-Bilyk & Goodman, 2004; Goodman et al., 2000). The DAWBA interview was administered by trained psychologists to primary caregivers when children were aged 6, 11 and 15 years. We used the DAWBA bands, a 6 category ordinal variable, derived from symptoms using a computerised algorithm, which represents the child’s probability of each condition being present (<0.1% (band 0), 0.5% (band 1), 3% (band 2), 15% (band 3), 50% (band 4), >70% (band 5)) (A. Goodman et al., 2011). Bands 4 and 5 are criteria for a likely diagnosis of a condition. Depression was assessed using the bands available. An “any anxiety condition” DAWBA band variable was derived using DAWBA bands for four anxiety conditions: separation anxiety, specific phobia, social anxiety, and generalised anxiety. For each individual, the highest likelihood band across these four conditions was used to define their likelihood of having any anxiety condition.

### Early-life risk factors

Early-life risk factors were grouped into the following three categories (summarised in Table 1):

**Table 1.** Summary of early risk factors included in the analysis.

| Early life-risk factors | Indicator | Measure | Time point of assessment | Coding |
| --- | --- | --- | --- | --- |
| **Socioeconomic** | Maternal education | Years of schooling | Birth | <9 or ≥ 9 years |
|  | Wealth index | Ownership of: vacuum cleaner; dishwasher; VCR/DVD; Refrigerator; Freezer/fridge-freezer; Microwave oven; Microcomputer; Landline telephone; Radio; Black and white TV; Colour TV; Car; Air conditioning unit; Presence of a maid | Birth | Quintiles (lowest versus rest) |
|  | Bolsa Família* recipient | Household receives Bolsa Família | Age 6 | Yes/No |
| **Maternal** | Support from child's father | Perceived support that mother received from the baby's father during the pregnancy | Birth | No or low/Some or high |
|  | Maternal depression | Edinburgh Postnatal Depression Scale | Age 1 | Yes/No; cut off ≥13 |
|  | Maternal age | Years | Birth | Continuous |
|  | Skin colour | Interviewer-assessed | Birth | White/non-White |
|  | Cohabiting with partner | Mother lives with husband or partner | Birth | Yes/No |
| **Child** | Childhood emotional/behavioural problems | Child Behaviour Checklist (CBCL) | Age 4 | Continuous |
|  | Experienced trauma | DAWBA PTSD items | Age 6 | Yes/No |

\* Bolsa Família is Brazil's major federal social welfare program, giving direct cash payments to low-income families, with the aim of fighting poverty, hunger, and social inequality (Neves et al., 2022).

#### 1. Socioeconomic factors

At birth, two socioeconomic indicators were assessed: i) maternal years of schooling, categorised as 0–8, or ≥ 9 years (8 years of education was compulsory in Brazil in 2004, equivalent to elementary education), and ii) household wealth index, constructed from ownership of several assets that was derived into quintiles, then binarised using the lowest wealth quintile (poorest) versus the other four higher wealth quintiles. From the age 6 follow-up, current receipt of Bolsa Família (a conditional cash transfer programme targeted at households living in poverty) was assessed and defined as a binary variable.

#### 2. Maternal factors

Maternal age (in years), mother-reported support from child’s father (“How did you feel about the support you received from the baby’s father during your pregnancy?”; A lot/some support vs little/no support), and whether the mother was cohabiting with a partner, were all assessed at birth. Maternal skin colour was assessed by the interviewer at birth and grouped into White and non-White

Maternal mental health was assessed using the Edinburgh Postnatal Depression Scale when offspring were 12 months of age (Cox et al., 1987). We used a cut-off (≥13) validated in Pelotas to define depression (Matijasevich et al., 2014).

#### 3. Child factors

The Child Behaviour Checklist (CBCL), used to assess emotional and behavioural problems, was completed by mothers when offspring were 4 years of age (Achenbach, 1991). The CBCL is a 113-item questionnaire, rated on a 3-point Likert scale ranging from 0 (absent) to 2 (occurs often). It was operationalised continuously, with higher scores representing greater emotional/behavioural problems. The tool has been validated in a Brazilian population (Bordin et al., 1995).

To assess childhood trauma, we used the post-traumatic stress section of the DAWBA, completed when children were 6 years of age. This was delivered by a trained psychologist to the primary caregiver to determine retrospective exposure to any of the following traumas: attack or threat; physical abuse; sexual abuse; witnessed domestic violence; witnessed an attack, accident or sudden death; in a serious accident; fire or other disaster; or other trauma. Trauma was dichotomised as either present or absent.

### Statistical analysis

Results are reported following the Guidelines for Reporting on Latent Trajectory Studies (GRoLTs) checklist (van de Schoot et al., 2017). In-line with previous work in this sample, we used parallel-process latent-class growth analysis (LCGA) for ordinal categorical data to examine co-developmental trajectories of anxiety and depression (using DAWBA bands) (Shakeshaft et al., 2026). Our primary sample included participants who had complete anxiety or depression data for at least one timepoint (age 6, 11 or 15 years). Further details on how trajectories were derived are provided in Supporting Information.

To examine associations between early-life factors and trajectory classes, we used the manual bias-adjusted three step method (Heron et al., 2015; Vermunt, 2010; Wickrama et al., 2021). This accounts for uncertainties in class assignment when determining the strength of association. Sex was included as a covariate in all analyses. The class with the lowest probability of anxiety and depression across development (*stable-low*) was used as the reference category. Post-hoc, as a secondary analysis to further assess the specificity of risk factors within elevated classes, we refitted our model to use the *decreasing* and *increasing* class as the reference category, which allowed comparisons between those with persistent-high likelihood of diagnosis compared to those with changing likelihood of disorder across childhood and adolescence. Odds ratios (OR) and 95% confidence intervals (CI) are presented. Analyses were conducted in MPlus (version 8.11). MPlus code can be found in Supporting Information.

### Missing data

Details of missing data for each variable can be found in Table S1. Firstly, for deriving trajectories, we used full information maximum likelihood to account for missing DAWBA band data, whereby any participant with DAWBA data for at least one condition at one timepoint was included. Secondly, to account for missing data in risk factor variables, we used multiple imputation by chained equations, using the R package ‘MICE’. The sample to be imputed was the same as for the trajectories (those with complete anxiety or depression data for at least one timepoint). As well as all analysis variables, auxiliary variables which predict missingness in the sample and/or predictive of the incomplete variables themselves were also included in the imputation model. These were maternal smoking during pregnancy, household income, total child strengths and difficulties questionnaire (SDQ) scores at ages 6, 11, 15 and 18, and depression and anxiety diagnoses at age 18. Variables were imputed using predictive mean matching using 5 nearest neighbours. 100 datasets were imputed with a burn-in of 25 iterations. Further detail on missing data is available in Supporting Information.

#### Sensitivity analyses

To assess the robustness of findings to missing data assumptions, we examined the associations between socioeconomic, maternal and child factors and anxiety and depression co-developmental trajectories using complete case (n=3,228) and observed risk factor variables. This was conducted for the primary analysis, which used the stable-low class as the reference group.

## Results

The sample included 3,815 participants with complete DAWBA data from at least one timepoint. The imputed and observed prevalence of anxiety and depression diagnoses, as well as early-life risk factors, are presented in Table 2. Frequency distributions of DAWBA band variables are presented in Table S2. Sex-stratified prevalences of anxiety and depression diagnoses are presented in Table S3.

**Table 2.** Risk factor and mental health summary statistics (imputed and observed data)

| Variable | Imputed<br>(n=3,815) | Observed* |
| --- | --- | --- |
|  | % / Mean (SE) |  |
| Female | 48.0% | 48.0% |
| **Risk factor** |  |  |
| *Socioeconomic* |  |  |
| <9 years maternal schooling | 56.4% | 56.7% |
| Low wealth index | 23.2% | 23.3% |
| Bolsa familia recipient | 31.8% | 31.7% |
| *Maternal* |  |  |
| Maternal age (years) | 26.1 (0.1) | 26.2 (0.1) |
| Maternal skin colour (non-White) | 26.9% | 26.8% |
| Low/no support from child's father during pregnancy | 7.2% | 6.9% |
| Mother lives with partner | 84.2% | 84.2% |
| Maternal depression | 15.0% | 14.9% |
| *Child* |  |  |
| Child emotional/behavioural problems | 34.8 (0.3) | 34.7 (0.3) |
| Experienced trauma before age 6 | 12.4% | 12.4% |
| **DAWBA diagnoses** |  |  |
| Depression |  |  |
| Age 6 | 0.5% | 0.4% |
| Age 11 | 0.4% | 0.4% |
| Age 15 | 1.5% | 1.5% |
| Anxiety (any) |  |  |
| Age 6 | 2.2% | 2.1% |
| Age 11 | 3.6% | 3.2% |
| Age 15 | 2.0% | 1.8% |
| Co-occurrence of anxiety and depression |  |  |
| Age 6 | 0.3% | 0.3% |
| Age 11 | 0.2% | 0.2% |
| Age 15 | 0.2% | 0.3% |

\*number of participants with observed data differs for each variable. Amount of missingness can be seen in Table S1.

### Anxiety and depression co-developmental trajectories

Table S4 shows model fit indices for parallel process LCGA models for anxiety and depression with one to five classes. The 4-class solution showed the best fit. As demonstrated previously and shown in Figure 1, the four classes can be characterised by: 1) *Stable-low likelihood of anxiety and depression diagnosis across childhood and adolescence* (53.9%), 2) *Increasing likelihood of diagnosis* (28.1%), 3) *Decreasing likelihood of diagnosis* (15.1%), and 4) *Persistent-high likelihood of diagnosis* (2.9%) (Shakeshaft et al., 2026).

**Figure 1.**
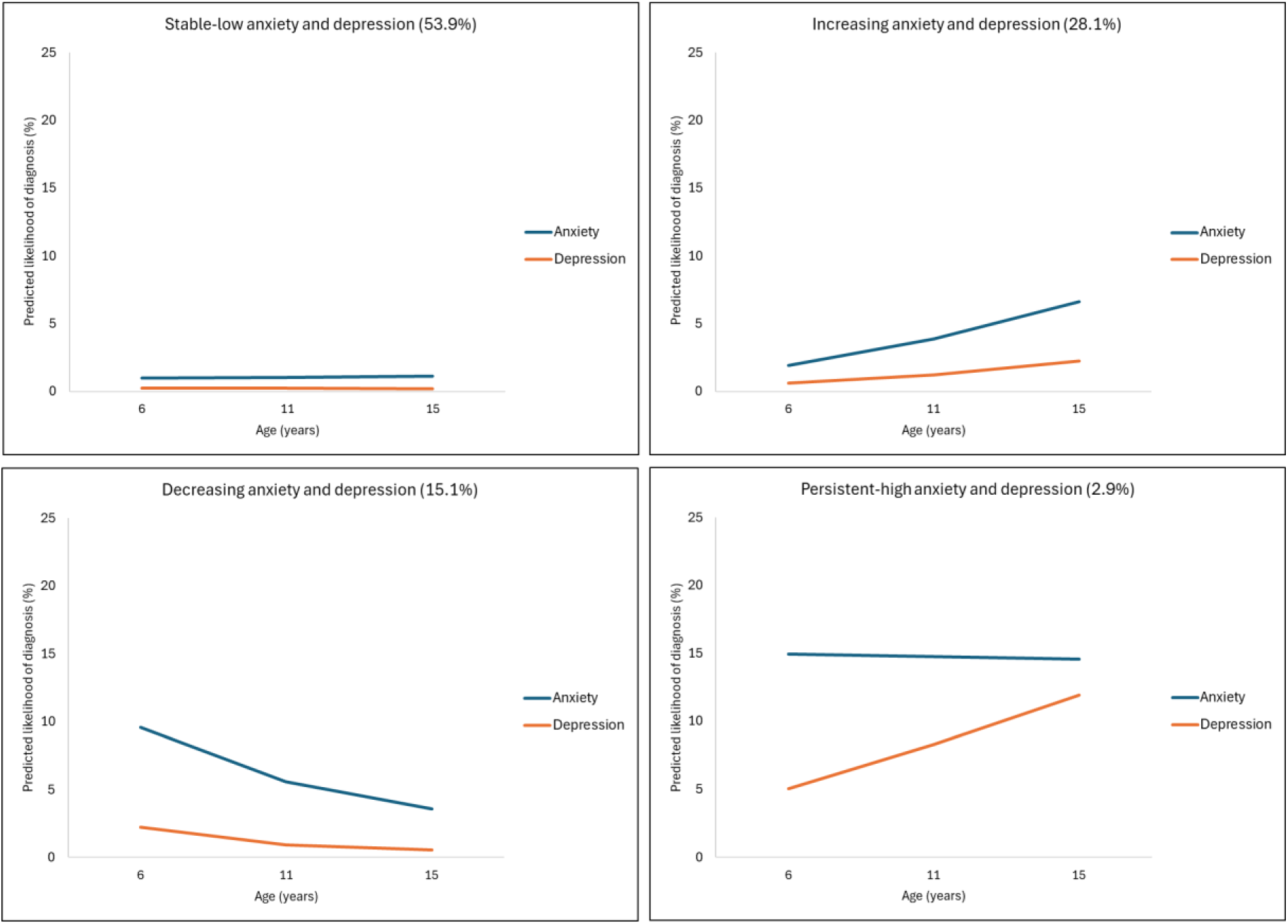
4-class latent-class growth analysis of anxiety and depression co-developmental trajectories.

### Early-life risk factors of anxiety and depression co-development

Table 3 shows associations between early-life factors and anxiety and depression co-development trajectory classes.

**Table 3.** Results of bias adjusted 3-step association between early-life factors and trajectory classes.

| Early-life factors | Trajectories of anxiety and depression co-development |  |  |  |
| --- | --- | --- | --- | --- |
|  | *Stable-low* | *Increasing* | *Decreasing* | *Persistent-high* |
| **Socioeconomic** |  |  |  |  |
| <9 years maternal education at birth | 1.00 (Ref) | 1.19 (0.88, 1.61) | 1.32 (0.94, 1.84) | 2.90 (1.28, 6.54) |
| Low wealth index at birth | 1.00 (Ref) | 1.22 (0.86, 1.73) | 1.19 (0.81, 1.74) | 1.36 (0.66, 2.82) |
| Bolsa Família recipient at age 6 | 1.00 (Ref) | 1.31 (0.95, 1.82) | 1.12 (0.78, 1.60) | 1.54 (0.81, 2.92) |
| **Maternal** |  |  |  |  |
| Age at birth<sup>a</sup> | 1.00 (Ref) | 0.98 (0.96, 1.00) | 0.97 (0.95, 0.99) | 0.98 (0.94, 1.03) |
| Skin colour (non-White) | 1.00 (Ref) | 1.24 (0.89, 1.73) | 1.25 (0.87, 1.79) | 1.53 (0.79, 2.95) |
| Low/no support from child's father during pregnancy | 1.00 (Ref) | 1.39 (0.77, 2.50) | 1.51 (0.83, 2.74) | 2.77 (1.20, 6.42) |
| Lives with partner at birth | 1.00 (Ref) | 0.65 (0.44, 0.96) | 0.77 (0.49, 1.21) | 0.52 (0.25, 1.10) |
| Maternal depression at age 1 | 1.00 (Ref) | 2.52 (1.58, 4.04) | 4.54 (2.98, 6.93) | 5.49 (2.77, 10.88) |
| **Child** |  |  |  |  |
| Emotional and behavioural problems at age 4<sup>a</sup> | 1.00 (Ref) | 1.06 (1.05, 1.07) | 1.09 (1.07, 1.10) | 1.10 (1.08, 1.12) |
| Experienced trauma before age 6 | 1.00 (Ref) | 1.09 (0.61, 1.96) | 3.62 (2.44, 5.37) | 3.21 (1.51, 6.85) |

All analyses are adjusted for sex. Odds ratios and 95% confidence intervals are shown for all associations. <sup>a</sup> = continuous variable. Trajectories are characterised by: 1) *Stable-low anxiety and depression across development*, 2) *Increasing anxiety and depression*, 3) *Decreasing anxiety and depression*, and 4) *Persistent-high anxiety and depression*.

#### Socioeconomic factors

There was strong evidence that lower maternal education at the time of their child’s birth was associated with persistent-high likelihood of anxiety and depression diagnosis (OR: 2.90, 95% CI: 1.28, 6.54) across childhood and adolescence (Table 3).

#### Maternal factors

Older maternal age was associated with lower odds of decreasing (OR: 0.97, 95% CI: 0.95, 0.99), increasing (OR: 0.98, 95% CI: 0.96, 1.00), and persistent-high (OR: 0.98, 95% CI: 0.94, 1.03) likelihood of anxiety and depression diagnosis, though some CI crossed the null. There was strong evidence that lower support from the child’s father during pregnancy was associated with a persistent-high likelihood of anxiety and depression diagnosis (OR: 2.77, 95% CI: 1.20, 6.42). There was evidence to suggest that maternal cohabitation with a partner was associated with lower odds of increasing likelihood of anxiety and depression diagnosis (OR: 0.65, 95% CI: 0.44, 0.96). Maternal depression when the child was aged one was strongly associated with elevated odds of persistent-high (OR: 5.49, 95% CI: 2.77, 10.88), decreasing (OR: 4.54, 95% CI: 2.98, 6.93), and increasing (OR: 2.52, 95% CI: 1.58, 4.04) likelihood of anxiety and depression diagnoses.

#### Child factors

Higher early childhood emotional/behavioural problems were associated with increasing (OR: 1.06, 95% CI: 1.05, 1.07), decreasing (OR: 1.09, 95% CI: 1.07, 1.10) and persistent-high (OR: 1.10, 95% CI: 1.08, 1.12) likelihood of anxiety and depression diagnosis. Experiencing trauma before age 6 was strongly associated with decreasing (OR: 3.62, 95% CI: 2.44, 5.37) and persistent-high (OR: 3.21, 95% CI: 1.51, 6.85) likelihood of anxiety and depression diagnosis, whilst CIs for the increasing group crossed the null (OR: 1.09, 95% CI: 0.61, 1.96).

Post-hoc analyses were conducted to investigate early-life factors associated with differences between the three elevated anxiety and depression classes. Compared with children who had decreasing likelihood of diagnosis, children who had increasing likelihood had a lower likelihood of having a mother with depression at age 1 (OR: 0.56, 95% CI: 0.32, 0.95), lower levels of childhood emotional/behavioural problems (OR: 0.98, 95% CI: 0.96, 0.99), and were less likely to have experienced childhood trauma (OR: 0.30, 95% CI: 0.15, 0.59, Table S5). There was no strong evidence of differences between the decreasing and persistent-high likelihood of diagnoses in relation to the risk factors.

Compared with children who had increasing likelihood of diagnosis, children with persistent-high likelihood were more likely to have mothers with lower educational attainment (OR: 2.44, 95% CI: 1.03, 5.76) and depression (OR: 2.17, 95% CI: 1.03, 4.61), have higher levels of childhood emotional/behavioural problems (OR: 1.04, 95% CI: 1.02, 1.06), and to have experienced childhood trauma (OR: 2.94, 95% CI:1.17, 7.38, Table S6).

### Sensitivity analysis

Results from analyses using complete cases (n = 3,228) and all observed data (see Ns in Table S1) were similar to the main analysis using imputed data (Tables S7-9). Some exceptions in the complete case analysis were: 1) evidence of associations between lower wealth index and Bolsa Família receipt with an increasing likelihood of anxiety and depression compared to the stable-low class, which was not observed in the main analysis, 2) there was no longer evidence of an association between mothers cohabiting with their partner and reduced odds of being in the increasing class compared to the stable-low class, due to wider CI.

## Discussion

This study explored how early-life socioeconomic, maternal and child factors were related to the co-development of anxiety and depression across childhood and adolescence, using a prospective longitudinal Brazilian birth cohort. We found evidence that lower maternal education, low or no support from the child’s father during pregnancy, maternal depression, early emotional/behavioural problems, and experiencing early-life trauma were associated with a persistent-high likelihood of anxiety and depression diagnoses across childhood and adolescence. We also found evidence indicating that maternal depression was associated with all three elevated trajectory classes, while there was evidence that mothers cohabiting with their partner/husbands appeared less likely to have children whose anxiety and depression onset occurred during adolescence. Post-hoc analyses suggested these factors did not differentiate between children with a persistent-high likelihood of anxiety and depression diagnosis, and children whose likelihood decreases across childhood and adolescence. Although many of these factors have previously been associated with anxiety-only and depression-only trajectories in HICs, there is very little existing evidence from LMICs, nor on risk factors of co-development in either setting. While some of the factors identified may causally influence trajectories of mental health in childhood and adolescence, this cannot be ascertained from these results alone as residual confounding is possible.

Our findings suggest that early-life family and psychosocial factors may contribute to differences in child and adolescent mental health trajectories. In particular, maternal depression, which is a known risk factor for offspring anxiety and depression, was associated with trajectories of anxiety and depression development that were not stable-low, similar to findings investigating co-development from the Netherlands in a younger HIC population (de Lijster et al., 2019; S. H. Goodman et al., 2011; Matijasevich et al., 2015; Tirumalaraju et al., 2020; Tusa et al., 2025). Maternal depression may be associated with worse child mental health trajectories in LMICs through multiple interacting pathways, particularly via the early caregiving environment and parenting behaviours (e.g. harsh punishment) as well as broader familial socioeconomic adversity (Herba et al., 2016). Community-based home-visiting programmes aimed at improving support to the mother after birth have shown some success in improving maternal mental health in LMICs (Bliznashka et al., 2021; Kang et al., 2025; Rahman et al., 2008), including the Happy Child Programme in Brazil (Biete et al., 2024). Further work should examine how these interventions in turn influence child outcomes, including their mental health.

Comparisons between increasing and decreasing likelihood of diagnoses indicate that early maternal and child psychosocial factors, including maternal depression, child emotional/behavioural problems and experiencing trauma, were more strongly associated with decreasing likelihood than increasing likelihood. This suggests that these factors may characterise children with early-onset problems, whereas children whose likelihood of diagnosis increases during adolescence may be influenced by factors not captured by early-life measures. Although some of these psychosocial factors were associated with all three elevated trajectories, the stronger associations with persistent-high and decreasing classes suggest these factors may represent vulnerability associated with the early-onset of anxiety and depression in childhood. To better understand persistence of anxiety and depression throughout childhood and adolescence, we attempted to identify factors that distinguish persistence of symptoms from decreasing symptoms in children who had an elevated likelihood of diagnoses in early-childhood. Comparisons between these two groups (persistent-high and decreasing) provided little evidence that the factors assessed could distinguish between them. This may indicate that contemporaneous or recent factors e.g., bullying or adverse childhood experiences, may contribute more strongly to whether early problems decrease or persist than early-life indicators (Shanahan et al., 2011).

Socioeconomic factors have previously been shown to strongly predict mental health (Lund et al., 2010). However, in this study, only low maternal education was strongly associated with a persistent-high likelihood of anxiety and depression diagnoses (the most severe class). Lower maternal education likely reflects both reduced economic and knowledge-based resources, influencing health beliefs and behaviours, as well as shaping access to healthcare, future employment and parenting resources over the lifespan, which may increase risk of persistent-high likelihood of anxiety and depression diagnosis in offspring (Howe et al., 2012). We saw limited evidence of an association with increasing or decreasing likelihood of diagnoses of anxiety and depression, possibly suggesting these are driven by emerging stressors across development, rather than long-term socioeconomic conditions. There was not strong evidence that other socioeconomic indicators were associated with anxiety and depression co-development. This contrasts with previous LMIC research showing household income, but not parental education, was associated with internalising and externalising problems in young people (Lansford et al., 2018). Differences may reflect variation in socioeconomic indicators, as income may capture acute financial strain more directly than asset-based measures or Bolsa Família receipt. However, this should not be interpreted as evidence that socioeconomic factors are less important than family or psychosocial factors for child mental health. The socioeconomic indicators available in this study may not fully capture dimensions of material hardship that are most relevant to child mental health, such as income insecurity, food insecurity, debt, or exposure to economic shocks.

This study is one of the first to explore risk factors of anxiety and depression co-developmental trajectories in an LMIC setting, and a strength is the range of risk factors explored and prospective measurements from birth to adolescence. However, it is not without limitations. Firstly, we were unable to explore paternal factors due to a lack of data collected on these. Previous studies indicate that paternal factors may be associated with child mental health, directly, or, indirectly via maternal behaviours or mental health (Jansen et al., 2024). Furthermore, despite the use of multiple imputation to account for the varying levels of missing data in early-life risk factors, this missing data may still introduce bias if unmeasured predictors of attrition exist. Next, since the number of cases with a high likelihood of a diagnosis of specific anxiety conditions was low, we were unable to explore trajectories and risk factors of the co-development of these anxiety conditions with depression. Evidence indicates that anxiety conditions emerge at different developmental periods, and show differing co-occurrence with depression, therefore risk factors of the co-development of specific anxiety conditions with depression may vary (Rapee et al., 2009; Solmi et al., 2022). Additionally, risk factors were assessed at a single time point, and the accumulation of exposures over time may also influence mental health trajectories. Finally, our findings demonstrate associations rather than causal relationships, and therefore, future research should examine whether these risk factors causally contribute to child/adolescent mental health in LMICs to further inform potentially effective interventions.

## Conclusion

This study adds to the very limited evidence base on risk factors of anxiety and depression co-development across childhood and adolescence in LMICs. Some factors (e.g., maternal depression) are associated with anxiety and depression regardless of age at onset whilst others (e.g., childhood trauma) appear to be more strongly associated with trajectories characterised by anxiety and depression at different developmental time points and persistence patterns. Given many socioeconomic, family and child factors are closely interconnected, a multi-domain approach to prevention is likely needed to reduce mental health problems in children and adolescents.

## AI Statement

The Authors declares that they have not used AI in any capacity that would require disclosure in accordance with Wiley’s AI Guidelines, nor in any capacity that would reasonably require discloser for editors, reviewers and readers to properly evaluate their research or manuscript. The authors confirm that AI has not been used for purposes including but NOT limited to drafting and editing, to generate substantial text or restructure arguments, nor in the research methodology. The authors take full responsibility for the accuracy of this statement.

## Acknowledgements

We thank all 2004 Pelotas Birth Cohort Study participants, families and research team. This article used data from the 2004 Pelotas Birth Cohort Study, conducted by the Postgraduate Program in Epidemiology at Universidade Federal de Pelotas, with the collaboration of the Brazilian Public Health Association (ABRASCO). The Wellcome Trust, WHO, National Support Program for Centers of Excellence (PRONEX), Brazilian National Research Council (CNPq), Brazilian Ministry of Health, and Children’s Pastorate supported previous phases of the study. The 11-year follow-up was supported by the Department of Science and Technology (DECIT) of the Brazilian Ministry of Health, CNPq and the Research Support Foundation of the State of São Paulo (FAPESP; grant number: 2014/13864-6). The 15-year follow-up was supported by the DECIT, CNPq, FAPESP (grant number: 2020/07730-8), the Research Support Foundation of the State of Rio Grande do Sul (FAPERGS), and the L’Oréal-Unesco-ABC Program for Women in Science in Brazil-2020. The 18-year follow-up was supported by DECIT, CNPq (grant number: 409224/2021-9), FAPESP (Grant n° <u>2023/12905-0</u>), FAPERGS, L’Oréal-Unesco-ABC Program for Women in Science in Brazil-2020, and the All for Health Institute. The present analyses were also funded FAPESP/UKRI/ESRC (grant n° 2023/12905-0). LB was funded by grant MR/W006308/1 for the GW4 BIOMED MRC DTP, awarded to the Universities of Bath, Bristol, Cardiff, and Exeter from the Medical Research Council (MRC)/UKRI. LR is supported by the Wolfson Centre for Young People’s Mental Health, established with support from the Wolfson Foundation. AS, BF, HJ, AGS, IPV, LDH, AM, LTR, and FR are supported by the Triple A Project, a Mental Health Award from Wellcome (309183/Z/24/Z). LTR, ISS, and AM (grant n°307079/2026-0) are supported by CNPq Research Scholarships. This study was supported by the NIHR Biomedical Research Centre at University Hospitals Bristol NHS Foundation Trust and the University of Bristol. DK is funded through the Elizabeth Blackwell Institute for Health Research at the University of Bristol which is supported by the Wellcome Trust (204813/Z/16/Z) and also by the National Institute for Health and Care Research Bristol Biomedical Research Centre (BRC-1215-20011). The views expressed are those of the author(s) and not necessarily those of the NHS, NIHR or the Department of Health and Social Care. The funders had no role in study design, data collection and analysis, decision to publish or preparation of the manuscript.

## Ethical Information

For Pelotas 2004, in all phases of the study, ethical approval was obtained from the Comissão de Ética para Análise de Projetos de Pesquisa do Hospital das Clínicas da Faculdade de Medicina da Universidade de São Paulo (CAPPesq-HCFMUSP) – perinatal and 3 month: date 07/07/2003, ID: 40601116; age 1 date: 09/03/2005, ID: 40601113; age 2 date: 09/03/2005, ID: 40600006; age 4 date: 13/04/2007, ID: 40600007; age 6 date: 31/08/2010, ID: OF 35/10; age 11 date: 26/11/2014, ID: 889.753; age 15 date: 04/09/19, ID: 3.554.667; age 18 date: 22/01/2022, ID: 5.210.484. Full informed consent was provided by parents or legal guardians. At ages 11, 15, and 18 years, adolescents signed an informed assent form.

## Data Availability

Applications to use the data can be made by contacting the researchers of the 2004 cohort (see http://www.epidemio-ufpel.org.br/site/content/faculty/ for a list of key faculty members) and completing the application form (http://www.epidemio-ufpel.org.br/site/content/studies/formularios.php). A list of administered questionnaires at each timepoint can be accessed online (http://www.epidemio-ufpel.org.br/site/content/coorte_2004-en/questionnaires.php).

