## Supplementary materials for "Associations between early-life socioeconomic, family and child factors, and anxiety and depression co-developmental trajectories in Brazil - analysis of the 2004 Pelotas Birth Cohort"

**Supplementary material**

### Multiple Imputation

We used multiple imputation to account for missing data in the cohort, with the assumption that data were Missing at Random (MAR). Proportions of missingness for all variables are shown in Table S1. Missingness was greatest in the mental health measures assessed at the age 15 follow-up (49.12%), mostly attributed to pauses in data collection during the COVID-19 pandemic. Missingness in the predictor variables ranged between 0.03% (maternal skin colour and cohabitation status) and 6.06% (childhood trauma). Around 17% of participants had missing data on mental health outcomes at age 18. Multiple imputation with chained equations was used to impute missing predictor and outcome data, under the MAR assumption. DAWBA measures of mental health used in the trajectories were imputed for summary statistics purposes only. Participants were included in the imputation if they had at least one anxiety or depression DAWBA measure at ages 6, 11 or 15.

*Table S1. Summary of missing data*

| Variable | Observed N | Missing N | Missing % |
|---|---|---|---|
| **Trajectory measures** | | | |
| Any anxiety (age 6) | 3,584 | 231 | 6.06 |
| Any anxiety (age 11) | 3,563 | 252 | 6.61 |
| Any anxiety (age 15) | 1,941 | 1,874 | 49.12 |
| Depression (age 6) | 3,584 | 231 | 6.06 |
| Depression (age 11) | 3,563 | 252 | 6.61 |
| Depression (age 15) | 1,941 | 1,874 | 49.12 |
| **Sociodemographic/Predictors** | | | |
| Sex (birth) | 3,815 | 0 | 0.00 |
| Maternal education (birth) | 3,779 | 36 | 0.94 |
| Wealth index (birth) | 3,810 | 5 | 0.13 |
| Bolsa Família recipient (age 6) | 3,610 | 205 | 5.37 |
| Support from child's father (pregnancy) | 3,771 | 44 | 1.15 |
| Maternal depression (age 1) | 3,618 | 197 | 5.16 |
| Maternal age (birth) | 3,812 | 3 | 0.08 |
| Maternal skin colour (birth) | 3,814 | 1 | 0.03 |
| Maternal cohabitation status (birth) | 3,814 | 1 | 0.03 |
| Child emotional/behavioural problems (age 4) | 3,627 | 188 | 4.93 |
| Childhood trauma (before age 6) | 3,584 | 231 | 6.06 |

Missing N and % based on number of individuals who have at least one anxiety or depression measurement at ages 6, 11 or 15.

**Trajectories derivation**

Parallel-process LGCA allows for the inclusion of more than one growth trajectory for different constructs simultaneously. In LCGA models, within class variability is set to zero thereby assuming that everyone within the same class follows the same trajectory. Our primary sample included participants who had complete anxiety or depression data for at least one timepoint (age 6, 11 or 15 years). We considered one to five different class solutions and selected the best fitting individual model based on model fit indices including log-likelihood (LL), Akaike's Information Criterion (AIC), Bayesian Information Criterion (BIC), Lo-Mendell-Rubin Likelihood Ratio Test (LMR-LMR) and Bootstrapped likelihood ratio test (LRT). Lower values of AIC/BIC suggest better model fit, whilst p-values <0.05 from the LMR/Bootstrapped LRT indicate additional classes improve model fit. We also considered the interpretability of groups and smallest class sizes (whereby classes with N <100 were not deemed acceptable). Once the optimal class number was determined, we re-ran the model with 500 random starts to ensure there were no issues with local maxima. Class sizes were based on the estimated model and reported to the nearest integer. As we had three time points, we investigated model fit for a linear function of time only.

*Table S2. Development and Well-Being Assessment (DAWBA) band summary statistics (imputed data)*

| | Age 6 | Age 11 | Age 15 |
|---|---|---|---|
| **Trajectory measures** | | | |
| *Anxiety bands* | | | |
| 0.5% | 79.0% | 73.2% | 71.5% |
| 3% | 14.3% | 17.4% | 22.5% |
| 15% | 4.6% | 6.2% | 4.3% |
| 50% | 1.2% | 2.4% | 1.5% |
| >70% | 0.9% | 0.8% | 0.3% |
| *Depression bands* | | | |
| <0.1% | 79.2% | 77.9% | 73.8% |
| 0.5% | 17.9% | 19.9% | 21.2% |
| 15% | 2.4% | 1.8% | 3.6% |
| 50% | 0.4% | 0.3% | 1.1% |
| >70% | 0.1% | 0.1% | 0.3% |

*Table S3. Sex-stratified mental health summary statistics (imputed data)*

| | Males | | | Females | | |
|---|---|---|---|---|---|---|
| | Age 6 | Age 11 | Age 15 | Age 6 | Age 11 | Age 15 |
| **Trajectory measures** | | | | | | |
| Depression diagnosis | 0.5% | 0.5% | 1.0% | 0.4% | 0.3% | 2.1% |
| Anxiety diagnosis | 2.3% | 3.5% | 1.6% | 2.1% | 3.7% | 2.4% |
| Specific phobia | 1.5% | 2.4% | 1.3% | 1.2% | 2.6% | 1.8% |
| Social anxiety | <0.3% | <0.3% | <0.3% | <0.3% | <0.3% | 0.5% |
| Generalised anxiety | - | <0.3% | - | - | <0.3% | <0.3% |
| Separation anxiety | 1.0% | 1.1% | <0.3% | 0.9% | 1.1% | 0.3% |

*Table S4. Model fit of latent-class growth analysis of anxiety and depression joint trajectory models with 1-5 classes*

| | 1 Class | 2 Class | 3 Class | 4 Class | 5 Class |
|---|---|---|---|---|---|
| **Parameters** | 10 | 15 | 20 | 25 | 30 |
| **Log-likelihood** | -12820 | -12315 | -12266 | -12223 | -12209 |
| **AIC** | 25659 | 24661 | 24573 | 24495 | 24478 |
| **BIC** | 25722 | 24755 | 24698 | 24652 | 24665 |
| **Entropy** | - | 0.539 | 0.484 | 0.544 | 0.597 |
| **LMR-LRT p value** | - | <0.001 | 0.0039 | 0.011 | 0.258 |
| **BLRT p value** | - | <0.001 | <0.001 | <0.001 | <0.001 |
| **Class 1 N (%)** | 3816 (100) | 2573 (67.4) | 941 (24.7) | 2056 (53.9) | 487 (12.8) |
| **Class 2 N (%)** | - | 1242 (32.6) | 2225 (58.3) | 1073 (28.1) | 1038 (27.2) |
| **Class 3 N (%)** | - | - | 649 (17.0) | 576 (15.1) | 2090 (54.8) |
| **Class 4 N (%)** | - | - | - | 110 (2.9) | 99 (2.6) |
| **Class 5 N (%)** | - | - | - | - | 102 (2.7) |

AIC, Akaike information criterion; BIC, Bayesian information criterion; BLRT, Bootstrap Likelihood Ratio Test; LMR-LRT, Lo–Mendell–Rubin adjusted likelihood ratio test

*Table S5. Results of bias adjusted 3-step association between early-life factors and trajectory classes, using decreasing as the reference category*

| **Early-life factors** | **Trajectories** | | |
|---|---|---|---|
| | *Increasing* | *Decreasing* | *Persistent-high* |
| **Socioeconomic** | | | |
| <9 years maternal education at birth | 0.90 (0.57, 1.44) | 1.00 (Ref) | 2.20 (0.87, 5.61) |
| Low wealth index at birth | 1.03 (0.61, 1.75) | 1.00 (Ref) | 1.15 (0.47, 2.78) |
| Bolsa Família recipient at age 6 | 1.18 (0.72, 1.93) | 1.00 (Ref) | 1.38 (0.63, 3.01) |
| **Maternal** | | | |
| Age at birth[a] | 1.01 (0.97, 1.04) | 1.00 (Ref) | 1.01 (0.96, 1.07) |
| Skin colour (non-White) | 1.00 (0.60, 1.64) | 1.00 (Ref) | 1.23 (0.55, 2.73) |
| Low/no support from child's father during pregnancy | 0.92 (0.41, 2.09) | 1.00 (Ref) | 1.84 (0.63, 5.42) |
| Lives with partner at birth | 0.84 (0.47, 1.52) | 1.00 (Ref) | 0.68 (0.27, 1.71) |
| Maternal depression at age 1 | 0.56 (0.32, 0.95) | 1.00 (Ref) | 1.21 (0.54, 2.70) |
| **Child** | | | |
| Emotional and behavioural problems at age 4[a] | 0.98 (0.96, 0.99) | 1.00 (Ref) | 1.02 (1.00, 1.03) |
| Experienced trauma before age 6 | 0.30 (0.15, 0.59) | 1.00 (Ref) | 0.89 (0.37, 2.14) |

All analyses are adjusted for sex. Odds ratios and 95% confidence intervals are shown for all associations. [a] = continuous variable.
Trajectories are characterised by: 1) *Stable-low anxiety and depression across development*, 2) *Increasing anxiety and depression*, 3) *Decreasing anxiety and depression*, and 4) *Persistent-high anxiety and depression. Findings for stable-low are not presented here as it was compared in the main analysis.*

*Table S6. Results of bias adjusted 3-step association between early-life factors and trajectory classes, using increasing as the reference category*

| **Early-life factors** | **Trajectories** | |
|---|---|---|
| | *Increasing* | *Persistent-high* |
| **Socioeconomic** | | |
| <9 years maternal education at birth | 1.00 (Ref) | 2.44 (1.03, 5.76) |
| Low wealth index at birth | 1.00 (Ref) | 1.11 (0.50, 2.46) |
| Bolsa Família recipient at age 6 | 1.00 (Ref) | 1.17 (0.58, 2.36) |
| **Maternal** | | |
| Age at birth[a] | 1.00 (Ref) | 0.99 (0.94, 1.05) |
| Skin colour (non-White) | 1.00 (Ref) | 1.23 (0.60, 2.54) |
| Low/no support from child's father during pregnancy | 1.00 (Ref) | 1.99 (0.76, 5.21) |
| Lives with partner at birth | 1.00 (Ref) | 0.81 (0.36, 1.80) |
| Maternal depression at age 1 | 1.00 (Ref) | 2.17 (1.03, 4.61) |
| **Child** | | |
| Emotional and behavioural problems at age 4[a] | 1.00 (Ref) | 1.04 (1.02, 1.06) |
| Experienced trauma before age 6 | 1.00 (Ref) | 2.94 (1.17, 7.38) |

All analyses are adjusted for sex. Odds ratios and 95% confidence intervals are shown for all associations. [a] = continuous variable.

Trajectories are characterised by: 1) *Stable-low anxiety and depression across development*, 2) *Increasing anxiety and depression*, 3) *Decreasing anxiety and depression*, and 4) *Persistent-high anxiety and depression. Findings for stable-low are not presented here as it was compared in the main analysis.*

### Sensitivity Analyses

*Table S7. Predictor summary statistics (imputed, observed and complete case data)*

| **Variable** | **Imputed N = 3815** | **Observed*** | **Complete case N = 3228** |
|---|---|---|---|
| | | %/ Mean (SE) | |
| Female | 48.0% | 48.0% | 48.0% |
| **Predictors** | | | |
| *Socioeconomic* | | | |
| <9 years maternal schooling | 56.4% | 56.7% | 56.3% |
| Low wealth index | 23.2% | 23.3% | 22.5% |
| Bolsa familia recipient | 31.8% | 31.7% | 32.2% |
| *Maternal* | | | |
| Maternal age (years) | 26.1 (0.1) | 26.2 (0.1) | 26.2 (0.1) |
| Maternal skin colour (non-White) | 26.9% | 26.8% | 26.7% |
| Low/no support from child's father during pregnancy | 7.2% | 6.9% | 6.7% |
| Mother lives with partner | 84.2% | 84.2% | 85.0% |
| Maternal depression | 15.0% | 14.9% | 14.8% |
| *Child* | | | |
| Child emotional/behavioural problems | 34.8 (0.3) | 34.7 (0.3) | 34.6 (0.3) |
| Experienced trauma before age 6 | 12.4% | 12.4% | 12.3% |

*Number of participants with observed data differs for each variable (can be seen in Table S1).

*Table S8. Results of bias adjusted 3-step association between early-life factors and trajectory classes, using complete case predictor variables (n = 3,228)*

| **Early-life factors** | **Trajectories** | | | |
|---|---|---|---|---|
| | *Stable-low* | *Increasing* | *Decreasing* | *Persistent-high* |
| **Socioeconomic** | | | | |
| <9 years maternal education at birth | 1.00 (Ref) | 1.33 (0.94, 1.88) | 1.12 (0.80, 1.58) | 2.76 (1.22, 6.22) |
| Low wealth index at birth | 1.00 (Ref) | 1.48 (1.00, 2.19) | 1.24 (0.82, 1.86) | 1.44 (0.68, 3.01) |
| Bolsa Família recipient at age 6 | 1.00 (Ref) | 1.46 (1.02, 2.08) | 1.03 (0.71, 1.50) | 1.62 (0.86, 3.06) |
| **Maternal** | | | | |
| Age at birth[a] | 1.00 (Ref) | 0.98 (0.96, 1.01) | 0.97 (0.95, 1.00) | 099 (0.94, 1.03) |
| Skin colour (non-White) | 1.00 (Ref) | 1.22 (0.83, 1.78) | 1.14 (0.78, 1.66) | 1.39 (0.71, 2.72) |
| Low/no support from child's father during pregnancy | 1.00 (Ref) | 1.10 (0.54, 2.23) | 1.33 (0.70, 2.51) | 2.74 (1.17, 6.40) |
| Lives with partner at birth | 1.00 (Ref) | 0.74 (0.47, 1.16) | 0.91 (0.56, 1.49) | 0.54 (0.26, 1.14) |
| Maternal depression at age 1 | 1.00 (Ref) | 2.74 (1.65, 4.55) | 4.00 (2.58, 6.20) | 6.16 (3.15, 12.05) |
| **Child** | | | | |
| Emotional and behavioural problems at age 4[a] | 1.00 (Ref) | 1.06 (1.05, 1,08) | 1.08 (1.07, 1.10) | 1.10 (1.08, 1.13) |
| Experienced trauma before age 6 | 1.00 (Ref) | 1.00 (0.52, 1.91) | 2.96 (1.95, 4.49) | 3.18 (1.53, 6.62) |

All analyses are adjusted for sex. Odds ratios and 95% confidence intervals are shown for all associations. [a] = continuous variable. Trajectories are characterised by: 1) *Stable-low anxiety and depression across development*, 2) *Increasing anxiety and depression*, 3) *Decreasing anxiety and depression*, and 4) *Persistent-high anxiety and depression*

*Table S9. Results of bias adjusted 3-step association between early-life factors and trajectory classes, using observed predictor variables*

| **Early-life factors** | **Trajectories** | | | |
|---|---|---|---|---|
| | *Stable-low* | *Increasing* | *Decreasing* | *Persistent-high* |
| **Socioeconomic** | | | | |
| <9 years maternal education at birth (n=3779) | 1.00 (Ref) | 1.19 (0.88, 1.61) | 1.29 (0.93, 1.79) | 2.84 (1.26, 6.36) |
| Low wealth index at birth (n=3810) | 1.00 (Ref) | 1.22 (0.86, 1.73) | 1.19 (0.81, 1.75) | 1.36 (0.65, 2.81) |
| Bolsa Família recipient at age 6 (n=3610) | 1.00 (Ref) | 1.36 (0.98, 1.89) | 1.10 (0.77, 1.56) | 1.55 (0.82, 2.91) |
| **Maternal** | | | | |
| Age at birth[a] (n=3812) | 1.00 (Ref) | 0.98 (0.96, 1.00) | 0.97 (0.95, 1.00) | 0.98 (0.94, 1.03) |
| Skin colour (non-White, n=3814) | 1.00 (Ref) | 1.24 (0.89, 1.73) | 1.25 (0.87, 1.79) | 1.53 (0.80, 2.95) |
| Low/no support from child's father during pregnancy (n=3771) | 1.00 (Ref) | 1.36 (0.75, 2.48) | 1.48 (0.81, 2.72) | 2.92 (1.28, 6.70) |
| Lives with partner at birth (n=3814) | 1.00 (Ref) | 0.65 (0.44, 0.96) | 0.77 (0.49, 1.21) | 0.53 (0.25, 1.10) |
| Maternal depression at age 1 (n=3618) | 1.00 (Ref) | 2.53 (1.58, 4.05) | 4.52 (2.96, 6.88) | 5.39 (2.74, 10.59) |
| **Child** | | | | |
| Emotional and behavioural problems at age 4[a] (n=3627) | 1.00 (Ref) | 1.06 (1.05, 1.07) | 1.09 (1.07, 1.10) | 1.10 (1.08, 1.13) |
| Experienced trauma before age 6 (n=3584) | 1.00 (Ref) | 0.99 (0.52, 1.91) | 3.50 (2.39, 5.13) | 3.13 (1.45, 6.73) |

All analyses are adjusted for sex. Odds ratios and 95% confidence intervals are shown for all associations. [a] = continuous variable. Trajectories are characterised by: 1) *Stable-low anxiety and depression across development*, 2) *Increasing anxiety and depression*, 3) *Decreasing anxiety and depression*, and 4) *Persistent-high anxiety and depression*

### Document of Transparent Changes

#### Transparent Changes

1. **Description of change:** *Explored maternal living status as a predictor.*
    a. **Rationale:** *Included after discussions with a Young Person's Advisory Group who suggested this may be important.*
    b. **Effect of change on study results:** *None expected.*
2. **Description of change:** *Used DAWBA bands instead of continuous DAWBA scores in trajectory modelling.*
    a. **Rationale:** *The way in which continuous DAWBA scores have previously been calculated were not applicable in this cohort, due to the way in which DAWBA questions were asked, and the inclusion of skip questions.*
    b. **Effect of change on study results:** *Whilst the findings are likely to be similar as we used the same measure, just presented differently, continuous scores may have allowed us to model unconstrained models which would allow for within-class variation.*
3. **Description of change:** *Generated overall anxiety measure using highest DAWBA band score instead of extracting individual-level factor scores to derive a continuous latent anxiety score*
    a. **Rationale for this change:** *As above, we were unable to use the continuous scores, and therefore, this was the best option.*
    b. **Effect of change on study results:** *Factor scores would have allowed us to explore the effect of sub-types in more detail, with information on whether subscales load onto a single anxiety factor. By creating it using the highest DAWBA band, we have assumed they are one latent factor but this may not be the case. Due to lack of variability within the bands, we could not explore subtype trajectories individually or with depression and therefore, this is a limitation of the study.*
4. **Description of change:** *Separate modelling of depression and anxiety trajectories was not included as an analysis*
    a. **Rationale for this change:** *The separate trajectories have not been explored in this analysis as the individual trajectories are not the focus of this paper, and are presented elsewhere.*
    b. **Effect of change on study results:** *None expected*

**MPlus Code**

1. **Parallel-process LCGA of anxiety and depression**

```
VARIABLE:
names = idalt20250916 any_anx_band6 any_anx_band11 any_anx_band15
      p1depband_06anos p1depband_11anos p1depband_15anos;
usevar = any_anx_band6 any_anx_band11 any_anx_band15
        p1depband_06anos p1depband_11anos p1depband_15anos;
 categorical = p1depband_06anos p1depband_11anos p1depband_15anos
             any_anx_band6 any_anx_band11 any_anx_band15;
missing =.;
class = c(4);
idvariable = idalt20250916;

ANALYSIS:
 TYPE = MIXTURE;
STARTS = 500 10;
LRTSTARTS = 0 0 400 80;
LINK = LOGIT;

MODEL:
%OVERALL%
I_D S_D | p1depband_06anos@0 p1depband_11anos@5 p1depband_15anos@9;
I_A S_A | any_anx_band6@0 any_anx_band11@5 any_anx_band15@9;
```

**2. Bias-adjusted 3-step association of predictors with trajectory classes**

VARIABLE: NAMES ARE asexo p1depband_06anos p1depband_11anos meducation2 assets2 ae01 ae02 ethnicity p1depband_15anos any_anx_band6 any_anx_band11 any_anx_band15 support mmh bf6 cbcltotal trauma jmini_anyanxiety_withouocd jmini_dep afumo fp1ebdtot p1ebdtot arendtot hp1ebdtot jpebdtot fidade gidade hidadeanos jidadeanosfinal p1sepabandd_06anos p1sepabandi_06anos p1genaband_06anos p1sophband_06anos p1spphband_06anos p1sepabandd_11anos p1sepabandi_11anos p1genaband_11anos p1sophband_11 p1spphband_11anos p1sepabandd_15anos p1sepabandi_15anos p1genaband_15anos p1sophband_15 p1spphband_15anos Class GAD_6_diagnosis phobia_6_diagnosis social_anx_6_diagnosis sep_anx _6_diagnosis_ICD MDD_6_diagnosis any_anx_6_diagnosis GAD_11_diagnosis phobia_11_diagnosis social_anx_11_ diagnosis sep_anx_11_diagnosis MDD_11_diagnosis any_anx_11_diagnosis GAD_15_diagnosis phobia_15_diagnosis social_anx_15_diagnosis sep_anx_15_diagnosis MDD_15_diagnosis any_anx_15_diagnosis Anx_Dep6 Anx_Dep11 Anx_Dep15;

```
!trauma used as example
  usevar = Class trauma asexo;
  nominal = Class;
  missing =.;
  class = c(4);

 ANALYSIS:
   TYPE = MIXTURE;
 STARTS = 500 20;

 MODEL:
 %OVERALL%
 c#1 c#2 c#3 on trauma asexo;

    %c#1%
       [Class#1@4.150 Class#2@2.265 Class#3@3.493];

       %c#2%
        [Class#1@2.280 Class#2@3.085 Class#3@1.955];

       %c#3%

       [Class#1@-0.835 Class#2@-0.907 Class#3@-4.481];
       %c#4%
        [Class#1@7.874 Class#2@6.317 Class#3@10.133];
```
